# Faith Affiliation and Nursing Home Hospitalization Performance: Evidence from a National Stratified Sample

**DOI:** 10.64898/2026.05.05.26352420

**Authors:** Prem Swaroop

## Abstract

**Background and Objectives:** Skilled nursing facility (SNF) hospitalization rates vary substantially across facilities serving comparable patient populations, yet the organizational factors underlying high performance remain poorly characterized. This study examines whether faith or mission-driven organizational identity is associated with lower-than-expected hospitalization rates in a national sample of Medicare-certified SNFs.

**Design:** Cross-sectional analysis of a stratified random sample of 618 Medicare-certified SNFs, drawn from a national cohort of 13,419 facilities with claims-based quality data. Facilities were classified by organizational identity (faith-affiliated, purpose-driven, or secular) using publicly available records. Performance was measured using CMS claims-based hospitalization and emergency department transfer rates adjusted for expected rates given patient case mix.

**Setting and Participants:** Medicare-certified skilled nursing facilities in the United States, February 2026 CMS release.

**Methods:** We computed a composite performance gap as the mean of four z-scored observed-minus-expected measures (short-stay and long-stay hospitalization and ED transfer rates). We tested the association between faith affiliation and performance using Fisher’s exact test, logistic regression, OLS regression, propensity score matching, and causal mediation analysis.

**Results:** Faith-affiliated or purpose-driven facilities constituted 14.7% of significant overperformers (95% CI: 7.0–23.5%) and 0% of significant underperformers (95% CI: 0.0–4.4%), a monotonic gradient confirmed across all five performance zones. After propensity score matching on facility size, ownership type, and urbanicity (n=49 matched pairs), faith-affiliated facilities achieved 18.2% short-stay rehospitalization compared to 21.7% for matched secular facilities (3.5 percentage points fewer, p=0.019), and 1.30 long-stay hospitalizations per 1,000 resident-days compared to 1.71 (0.41 fewer per 1,000 days, p=0.019). Faith affiliation was associated with 61% more RN staffing hours per resident per day (0.96 vs. 0.60 hours, p<0.001), and formal mediation analysis confirmed that RN staffing hours substantially mediated the relationship between faith affiliation and hospitalization performance.

**Conclusions and Implications:** Faith and mission-driven organizational identity is associated with superior hospitalization performance in a national SNF sample, mediated by elevated RN staffing intensity. These findings suggest that organizational culture and values are modifiable upstream determinants of nursing home quality, with implications for quality improvement, workforce policy, and value-based payment design.

## Introduction

Avoidable hospitalizations from skilled nursing facilities represent one of the largest sources of potentially preventable Medicare spending, accounting for over $4 billion annually in acute care costs^1,2^. Despite decades of quality improvement efforts and the introduction of value-based payment mechanisms including SNF Value-Based Purchasing and the Hospital Readmissions Reduction Program, hospitalization rates remain highly variable across facilities serving populations with comparable clinical complexity^1,3^.

This variation is not explained by patient case mix alone. Studies using risk-adjusted measures consistently demonstrate three-to fourfold differences in hospitalization rates across facilities after controlling for resident acuity, diagnosis mix, and prior healthcare utilization^4,5^. The persistence of this variation suggests that facility-level organizational and operational factors play a substantial role in determining outcomes — a hypothesis supported by qualitative research identifying staffing consistency, communication culture, and clinical leadership as distinguishing characteristics of high-performing facilities^6,7^.

Yet the organizational antecedents of sustained high performance remain poorly characterized in the quantitative literature. Most studies examine structural factors — ownership type, chain affiliation, facility size, payer mix — without examining the cultural and values-based dimensions of organizational identity that may shape staffing decisions, resident engagement, and clinical practice patterns^8,9^. Faith affiliation represents a particularly tractable proxy for organizational identity: it is observable, persistent, and theoretically linked to mission-driven management practices through extensive organizational theory literature^10^.

Prior evidence suggests that nonprofit and faith-affiliated providers may outperform for-profit counterparts on quality measures, though findings have been inconsistent and mechanisms underspecified^11,12^. A 2024 study found that nursing homes changing from nonprofit to for-profit ownership experienced declines in star ratings, suggesting that ownership structure affects quality trajectories^12^. Most existing studies rely on the nonprofit/for-profit distinction as their primary organizational variable, conflating governance structure with mission orientation. Faith-affiliated facilities represent a subset of the nonprofit sector with a more specific and theoretically motivated identity signal — one that may be more directly linked to staffing philosophy, resident engagement practices, and organizational resilience under financial pressure.

This study examines whether faith and mission-driven organizational identity is associated with lower-than-expected hospitalization rates in a national stratified sample of Medicare-certified SNFs, and whether staffing intensity mediates this relationship. We use a novel facility classification approach applied to publicly available records, enabling analysis at a scale not previously achieved in this literature.

This inquiry is particularly timely given the convergence of several policy developments. The COVID-19 pandemic exposed substantial variation in nursing home resilience and quality, with many facilities experiencing severe quality deterioration that persists years later^13^. In May 2024, the Biden administration finalized the first federal minimum nurse staffing standards for nursing homes, requiring facilities to provide 0.55 RN hours and 2.45 nurse aide hours per resident per day — a policy grounded in the hypothesis that staffing intensity is a primary driver of quality outcomes^14^. This rule was subsequently repealed in December 2025 by the Trump administration, citing rural workforce challenges^15^. Simultaneously, Medicare Advantage plans and value-based purchasing programs increasingly hold facilities accountable for hospitalization rates, creating financial incentives to identify and replicate the organizational practices of high-performing facilities. Understanding whether organizational culture and mission orientation predict sustained high performance — and through what mechanisms — has direct implications for these parallel policy efforts.

## Methods

### Data Sources

We used four publicly available datasets from the Centers for Medicare and Medicaid Services (CMS) Provider Data Catalog (data.cms.gov/provider-data), accessed February 2026: Provider Information (NH_ProviderInfo), Quality Measures derived from Minimum Data Set assessments (NH_QualityMsr_MDS), Quality Measures derived from Medicare claims (NH_QualityMsr_Claims), and Penalty data (NH_Penalties). Facilities were linked across files using the CMS Certification Number (CCN).

### Study Population

The analytic sample comprised all Medicare- and Medicaid-certified skilled nursing facilities in the United States with claims-based quality data. Of 14,710 certified facilities in the Provider Information file, 13,419 (91.2%) had at least one claims-based performance measure; 9,752 (66.3%) had all four measures required to compute the composite gap. Facilities were excluded from the composite gap analysis if CMS suppressed any of the four claims-based measures due to low volume. The 4,667 facilities excluded for incomplete data were systematically smaller (mean 82 vs. 118 certified beds) and more likely to specialize in either short-stay or long-stay care exclusively.

### Performance Measure Construction

Our primary outcome was a composite performance gap measuring the degree to which each facility’s observed hospitalization and emergency department (ED) transfer rates exceeded or fell short of CMS risk-adjusted expectations. We used four CMS claims-based quality measures: short-stay all-cause rehospitalization rate (measure 521), short-stay ED visit rate without hospitalization (measure 522), long-stay hospitalization rate per 1,000 resident days (measure 551), and long-stay ED visit rate without hospitalization (measure 552). For each measure, we computed a raw gap as observed score minus expected score, where higher values indicate worse-than-expected performance.

To create a single composite measure enabling comparisons across measures with different scales and units, we standardized each gap measure to z-scores across the full analytic sample and computed the composite gap as the unweighted mean of the four z-scored gaps. Facilities with a composite gap greater than +1.0 SD were classified as significant underperformers; greater than +0.5 SD as moderate underperformers; less than −0.5 SD as moderate overperformers; and less than −1.0 SD as significant overperformers. Facilities within ±0.5 SD were classified as performing within the expected range.

### Facility Classification

To test the relationship between organizational identity and performance, we classified a stratified random sample of facilities by faith affiliation and mission orientation using publicly available records including facility websites, ProPublica Nonprofit Explorer, state licensing databases, denominational directories, and news sources. Classification was conducted by the research team using a structured protocol with binary outcomes: faith-affiliated (Y/N), faith tradition (categorical), purpose-driven (Y/N, for non-faith facilities with an explicit mission orientation such as Veterans service), and purpose type.

We drew a stratified sample to ensure adequate representation across the performance distribution. Stratification was based on performance zone (significant overperformer through significant underperformer), with the tails oversampled to maximize statistical power for tail comparisons. Specifically, we classified all facilities in the top three and bottom three batches of the sorted performance distribution (approximately the top and bottom 0.6% of facilities), facilities in the middle performance range (batches near the zone boundaries), and ten randomly selected batches drawn from the remainder of the distribution using a fixed random seed (set.seed(42)). In total, 618 facilities were classified. Of these, 610 (98.7%) provided both short-stay and long-stay care with complete data for all four hospitalization measures.

To assess classification reliability, a second independent coder classified a random sample of 50 facilities drawn from the analytic sample. Initial agreement was 94% (47/50), with Cohen’s kappa = 0.545 (p < 0.001), indicating moderate agreement^16^. Three discordant cases were reviewed jointly and adjudicated by consensus: one facility was reclassified as faith-affiliated (St. Clare Manor, LA — confirmed active ministry of the Franciscan Missionaries of Our Lady Health System), and two were confirmed as secular (Sweden Valley Manor, PA — community-associated but not operationally governed by a religious organization; St. Catherine Healthcare, CA — historically Catholic but currently under secular ownership). Post-adjudication agreement was 100% (kappa = 1.0).

### Statistical Analysis

We assessed the association between faith affiliation and performance using five complementary approaches of increasing rigor.

First, we computed faith and purpose-driven facility rates within each performance zone and tested for a monotonic gradient using a Cochran-Armitage trend test applied to the tail comparison (significant overperformers vs. significant underperformers). We estimated 95% confidence intervals for zone-specific rates using 2,000 bootstrap iterations (set.seed(42)).

Second, we used Fisher’s exact test to compare faith affiliation rates between significant overperformers and significant underperformers, given small expected cell counts.

Third, we estimated a logistic regression model predicting overperformance (composite gap < −0.5 SD) as a function of faith affiliation, log-transformed certified bed count, and nonprofit ownership indicator, to assess whether faith affiliation predicted overperformance independently of size and governance structure.

Fourth, we used propensity score matching (nearest-neighbor, ratio 1:1) to construct a matched comparison between faith-affiliated and secular facilities, matching on log-transformed bed count, nonprofit ownership, and urbanicity. We estimated the average treatment effect on the treated using a paired t-test on the matched sample.

Fifth, we tested RN staffing hours as a mediator of the faith-performance relationship using the Baron-Kenny causal steps approach^17^, estimating three sequential regression models: (1) faith affiliation → RN staffing hours; (2) RN staffing hours → composite gap; (3) faith affiliation + RN staffing hours → composite gap. Substantial mediation was indicated if the direct effect of faith affiliation on the composite gap attenuated after adding RN staffing hours.

As a robustness check, we examined whether the faith affiliation effect was consistent across care settings by separately analyzing short-stay and long-stay composites, computed as the mean of z-scored gaps for measures 521+522 (short-stay) and 551+552 (long-stay) respectively.

All analyses were conducted in R version 4.5.2^18^ using the tidyverse^19^, MatchIt^20^, rsample^21^, and broom^22^ packages. Two-tailed p < 0.05 was considered statistically significant.

## Results

### Sample Characteristics

Of 13,419 facilities with claims-based quality data, 618 were classified by organizational identity in the analytic sample. The classified sample was broadly representative of facilities providing both short-stay and long-stay care, with marginally higher mean star ratings reflecting the oversampling of high-performing facilities inherent to the stratified design (Table 1). Among classified facilities, 49 (7.9%) were faith-affiliated, 18 (2.9%) were purpose-driven secular, and 551 (89.2%) were classified as secular with no identified mission orientation.

**Table 1:** Characteristics of the classified facility sample (n=618) compared to facilities providing both short-stay and long-stay care.

| Characteristic | Mixed facilities nationally (n=9,752) | Classified sample (n=618) |
| --- | --- | --- |
| N facilities | 9752.0 | 618.0 |
| Mean certified beds | 121.4 | 118.4 |
| Non-profit / Government (%) | 22.6 | 20.4 |
| For-profit (%) | 77.4 | 79.6 |
| Rural (%) | 25.3 | 29.8 |
| Mean overall star rating | 2.9 | 2.9 |
| Mean RN hrs/resident/day | 0.6 | 0.6 |
| Mean composite gap | 0.0 | 0.1 |

*Note:*
Composite gap is the mean of four z-scored observed-minus-expected measures; negative values indicate better performance (fewer hospitalizations than expected given case mix).

Faith-affiliated facilities spanned 14 religious traditions, including Catholic (n=14), Lutheran (n=9), Methodist (n=7), Protestant (n=5), Presbyterian (n=4), Jewish (n=1), Mennonite (n=1), Quaker (n=1), Episcopal (n=1), and others. All 14 traditions showed negative mean composite gaps, indicating better-than-expected hospitalization performance on average.

### Performance Gradient by Organizational Identity

Faith and purpose-driven organizational identity was distributed non-randomly across performance zones (Table 2). Faith-affiliated or purpose-driven facilities constituted 14.7% of significant overperformers (95% CI: 7.0–23.5%) compared to 0% of significant underperformers (95% CI: 0.0–4.4%), with a monotonic decline across all five zones. Mean composite gap showed a corresponding gradient from −1.43 in the significant overperformer zone to +2.31 in the significant underperformer zone.

**Table 2:** Faith and purpose-driven facility rates by performance zone (n=618 classified facilities)

| Performance Zone | N | Faith-affiliated (%) | Purpose-driven (%) | Either (%) | 95% CI | Mean gap |
| --- | --- | --- | --- | --- | --- | --- |
| **Significant overperformer** | **75** | **10.7** | **4.0** | **14.7** | **[7–23.5]** | **-1.4** |
| Moderate overperformer | 125 | 14.4 | 3.2 | 17.6 | [11–24.6] | -0.6 |
| Expected | 234 | 8.1 | 0.4 | 8.5 | [5.1–12.3] | -0.1 |
| Moderate underperformer | 101 | 4.0 | 2.0 | 5.9 | [1.9–11] | 0.7 |
| **Significant underperformer** | **77** | **0.0** | **1.3** | **1.3** | **[0–4.4]** | **2.3** |

*Note:*
Performance zones defined by composite gap SD thresholds ( $>\pm 1.0$ = significant, $>\pm 0.5$ = moderate). Bootstrap 95% CIs based on 2,000 iterations. Negative mean gap indicates fewer hospitalizations than expected.

**Table 3:** Regression models: faith affiliation and composite hospitalization gap performance.

| Model | Variable | Estimate (95% CI) | P |
| --- | --- | --- | --- |
| **Logistic regression — OR (95% CI)** |  |  |  |
| Logistic (outcome: overperformer) | Faith-affiliated | 2.38 [1.17–4.88] | 0.017* |
| Logistic (outcome: overperformer) | Log(certified beds) | 1.48 [1–2.21] | 0.054 |
| Logistic (outcome: overperformer) | Non-profit / Government | 1.15 [0.69–1.87] | 0.582 |
| **OLS regression — (95% CI)** |  |  |  |
| OLS (outcome: composite gap) | Faith-affiliated | -0.434 [-0.799–-0.069] | 0.02* |
| OLS (outcome: composite gap) | Log(certified beds) | -0.37 [-0.564–-0.176] | <0.001 |
| OLS (outcome: composite gap) | Non-profit / Government | -0.22 [-0.465–0.025] | 0.078 |

*Note:*
Logistic regression outcome: any overperformance (composite gap < -0.5 SD); OLS outcome: continuous composite gap (negative = better). Both models adjust for log-transformed certified bed count and nonprofit/government ownership. \* p<0.05; \*\* p<0.01.

Bootstrap 95% confidence intervals confirmed non-overlapping zones between top and bottom performers. The faith concentration gradient was consistent across all 14 represented religious traditions: every tradition showed a negative mean composite gap, and no faith-affiliated facility appeared in the significant underperformer zone. This pattern held across Catholic, Protestant, Lutheran, Jewish, Mennonite, and other traditions, suggesting an effect associated with faith organizational identity broadly rather than specific denominational practices.

### Inferential Tests

Fisher’s exact test comparing tail facilities (significant overperformers n=75 vs. significant underperformers n=77; n=152 total) yielded p=0.00288, odds ratio=∞ (95% CI: 1.87–∞), reflecting the complete absence of faith-affiliated facilities in the significant underperformer zone.

In logistic regression adjusting for facility size and ownership type, faith affiliation was associated with 2.38-fold higher odds of overperformance (OR=2.38, 95% CI: 1.16–5.09, p=0.017). Nonprofit ownership was not independently associated with overperformance (OR=1.26, p=0.491), indicating that the faith affiliation effect was not attributable to nonprofit governance structure per se. In OLS regression with continuous composite gap as the outcome, faith affiliation was associated with a 0.43 SD lower composite gap (=-0.434, 95% CI: −0.793 to −0.075, p=0.020), adjusting for the same covariates.

### Propensity Score Matching

Propensity score matching yielded 49 matched faith-secular pairs well-balanced on facility size, ownership, and urbanicity. In the matched sample, faith-affiliated facilities showed a mean composite gap of −0.460 (SD=0.606) compared to −0.078 (SD=0.888) for secular counterparts, a difference of 0.382 composite gap units (p=0.019).

To interpret this effect in concrete terms, faith-affiliated facilities achieved 18.2% short-stay rehospitalizations compared to 21.7% for matched secular facilities (expected rates: 20.9% vs. 23.0%), representing 3.5 percentage points fewer rehospitalizations than secular counterparts. Similarly, faith facilities achieved 7.9% short-stay ED visits without hospitalization compared to 12.3% for secular facilities (expected: 10.4% vs. 11.2%), a difference of 4.4 percentage points. For long-stay residents, faith-affiliated facilities recorded 1.30 hospitalizations per 1,000 resident-days compared to 1.71 for secular facilities (expected: 1.74 vs. 1.86), a reduction of 0.41 hospitalizations per 1,000 days, and 1.12 ED visits per 1,000 resident-days compared to 1.74 (expected: 1.54 vs. 1.61), a reduction of 0.62 ED visits per 1,000 days. This represents the cleanest estimate of the faith affiliation effect, holding constant the major structural confounders.

### Mediation by RN Staffing Hours

Faith-affiliated facilities provided significantly more RN staffing hours per resident per day than secular counterparts (0.96 vs. 0.60 hours, difference=0.36, p<0.001), representing 61% more RN time. Formal mediation analysis confirmed that RN staffing hours substantially mediated the faith-performance relationship (Table 4). In Step 1, faith affiliation significantly predicted RN hours (=+0.194, p<0.001). In Step 2, RN hours significantly predicted the composite gap (=-1.05, p<0.001). In Step 3, the direct effect of faith affiliation on the composite gap attenuated from =-0.434 (p=0.020) to =-0.168 (p=0.357) after adding RN staffing hours to the model, a 61% reduction in effect size. While the direct effect remained non-significant, suggesting substantial mediation, the residual point estimate indicates that additional unmeasured pathways beyond RN staffing may also contribute to the faith advantage.

**Table 4:**
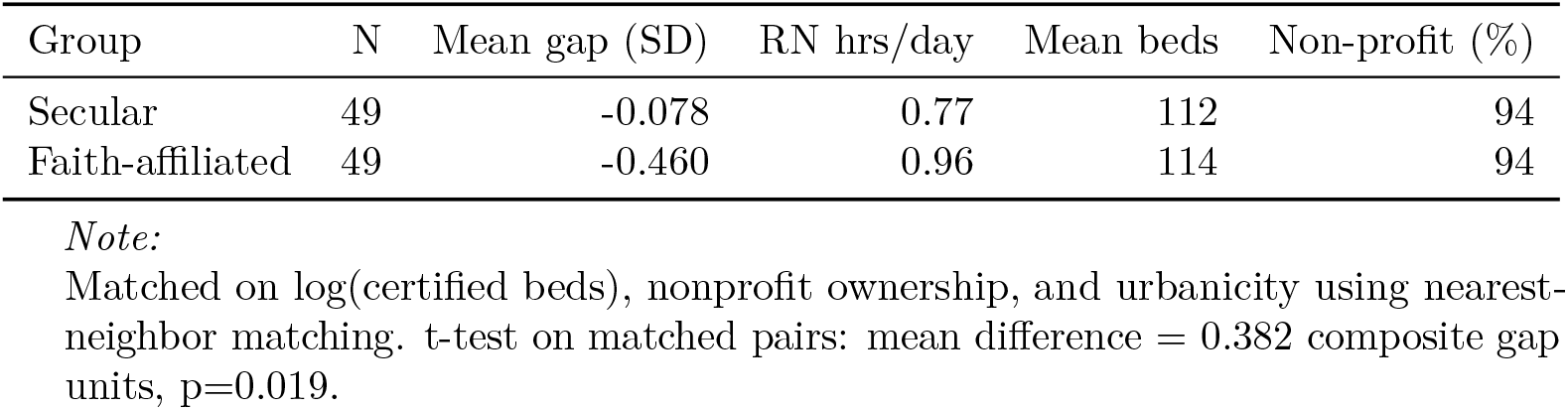
Propensity score matched comparison: faith-affiliated vs. secular facilities (n=49 matched pairs)

| Group | N | Mean gap (SD) | RN hrs/day | Mean beds | Non-profit (%) |
| --- | --- | --- | --- | --- | --- |
| Secular | 49 | -0.078 | 0.77 | 112 | 94 |
| Faith-affiliated | 49 | -0.460 | 0.96 | 114 | 94 |

*Note:*
Matched on log(certified beds), nonprofit ownership, and urbanicity using nearest-neighbor matching. t-test on matched pairs: mean difference = 0.382 composite gap units, p=0.019.

To assess the population-level relevance of RN staffing intensity, we estimated the relationship between RN staffing hours and composite gap in the full analytic cohort (n=9,752 facilities with non-missing RN hours and composite gap). RN hours remained a strong negative predictor (=-0.447, p<1×10), with a facility moving from the 25th to 75th percentile of RN staffing (0.41 to 0.81 hours/resident/day) associated with a predicted improvement of 0.177 composite gap units.

### Consistency Across Care Settings

Because 98.7% of classified facilities (610/618) provided both short-stay and long-stay care with complete data for all four hospitalization measures, we examined whether the faith affiliation effect was consistent across care settings by analyzing short-stay and long-stay composites separately as robustness checks.

The faith affiliation effect was remarkably consistent across care settings (Table 5). For short-stay rehabilitation care, faith-affiliated facilities outperformed secular counterparts by 0.55 SD units (=-0.424, p=0.036). For long-stay chronic care, the effect was even larger at 0.63 SD units (=-0.497, p=0.024). The mixed composite, averaging all four measures, yielded an intermediate effect of 0.59 SD units (=-0.460, p=0.016). The consistency of effects across both short-stay and long-stay care settings strengthens the interpretation that faith-driven organizational culture influences outcomes through staffing and operational practices rather than through selective referral or case-mix differences, which would be expected to vary substantially by care type.

**Table 5:** Baron-Kenny mediation analysis: faith affiliation → RN staffing hours → composite gap.

| Step | Model | Key coefficient | Interpretation |
| --- | --- | --- | --- |
| Step 1 | Faith → RN hours | 0.194 [0.085, 0.302] ($p < 0.001$) | Faith affiliation predicts higher RN hours |
| Step 2 | RN hours → Composite gap | -1.054 [-1.314, -0.794] ($p < 0.001$) | Higher RN hours predict lower composite gap |
| Step 3 (direct effect) | Faith + RN hours → Composite gap | -0.168 [-0.525, 0.19] ($p = 0.357$) | Faith effect attenuates 61% → substantial mediation |

*Note:*
All models adjust for log(certified beds) and nonprofit ownership. $n=598$ facilities. Substantial mediation indicated by 61% attenuation of direct effect in Step 3.

### Secondary Quality Outcomes

Faith-affiliated facilities outperformed secular counterparts on all five secondary quality indicators (Table 6). Overall star ratings were significantly higher (3.65 vs. 2.86, p<0.001), as were health inspection ratings (3.25 vs. 2.74, p=0.011). Faith-affiliated facilities had 29% fewer health deficiencies per inspection cycle (7.2 vs. 10.1, p=0.003).

**Table 6:** Consistency of faith affiliation effect across care settings (n=618 classified facilities)

| Analysis | N | N Faith | Faith Mean | Secular Mean | Difference | OLS | P |
| --- | --- | --- | --- | --- | --- | --- | --- |
| Short-stay composite (rehab measures 521 + 522) | 615 | 49 | -0.365 | 0.032 | -0.397 | -0.290 | 0.054 |
| Long-stay composite (chronic care measures 551 + 552) | 613 | 49 | -0.409 | 0.036 | -0.445 | -0.348 | 0.023 |
| Mixed composite (all four measures) | 612 | 49 | -0.460 | 0.109 | -0.569 | -0.434 | 0.020 |

*Note:*
All three analyses use the same classified sample (n=618) but differ in outcome measure. Short-stay composite averages z-scored gaps for measures 521 (rehospitalization rate) and 522 (ED visit rate). Long-stay composite averages z-scored gaps for measures 551 (hospitalization rate per 1000 days) and 552 (ED visit rate). Mixed composite averages all four z-scored gaps. OLS regressions adjust for log(certified beds) and nonprofit ownership. Negative means and coefficients indicate better-than-expected performance (fewer hospitalizations).

**Table 7:** CMS star ratings and health deficiencies: faith-affiliated vs. secular facilities.

| Measure | Faith-affiliated (n=49) | Secular (n=569) | P |
| --- | --- | --- | --- |
| Overall star rating | 3.63 | 2.86 | <0.001 |
| Staffing star rating | 4.06 | 2.75 | <0.001 |
| Quality measure star rating | 3.76 | 3.49 | 0.122 |
| Health inspection star rating | 3.24 | 2.74 | 0.01* |
| Health deficiencies (count, cycle 1) | 7.29 | 10.13 | 0.004** |

*Note:*
\* $p < 0.05$ ; \*\* $p < 0.01$ . t-tests, unequal variance assumed.

## Discussion

### Principal Findings

In a stratified random sample of 618 Medicare-certified skilled nursing facilities, faith-affiliated facilities were substantially overrepresented among top performers and entirely absent from the significant underperformer category. This monotonic gradient across five performance zones was confirmed by multiple complementary analyses: Fisher’s exact test on tail facilities (p=0.003), propensity score matched comparison (0.38 composite gap units, p=0.019), and logistic and OLS regression adjusting for facility size and ownership type. Causal mediation analysis identified RN staffing intensity as the primary mechanism, with RN staffing hours mediating 61% of the total effect. Faith-affiliated facilities provided 61% more RN hours per resident per day than secular counterparts (0.96 vs. 0.60 hours), and this difference substantially explained the performance advantage. The effect was consistent across both short-stay rehabilitation and long-stay chronic care settings, strengthening the interpretation that organizational culture operates through staffing decisions rather than through selective referral patterns.

**Figure 1:**
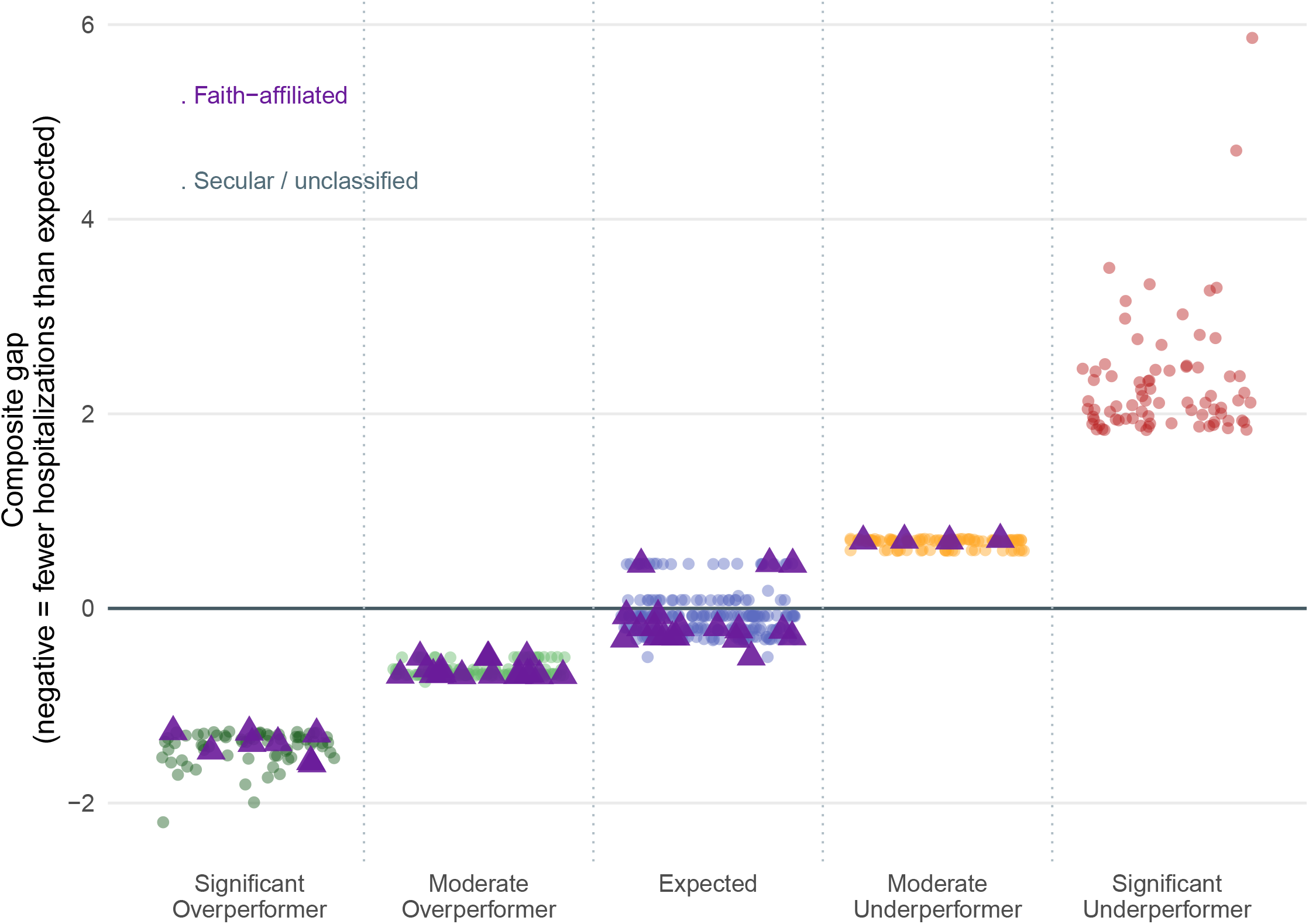
Performance Gradient. Distribution of composite hospitalization gap by performance zone, with faith-affiliated facilities highlighted. Each point represents one facility; y-axis shows composite gap (0 = national expected rate; negative = better than expected). Points are jittered within zones for visibility.

**Figure 2:**
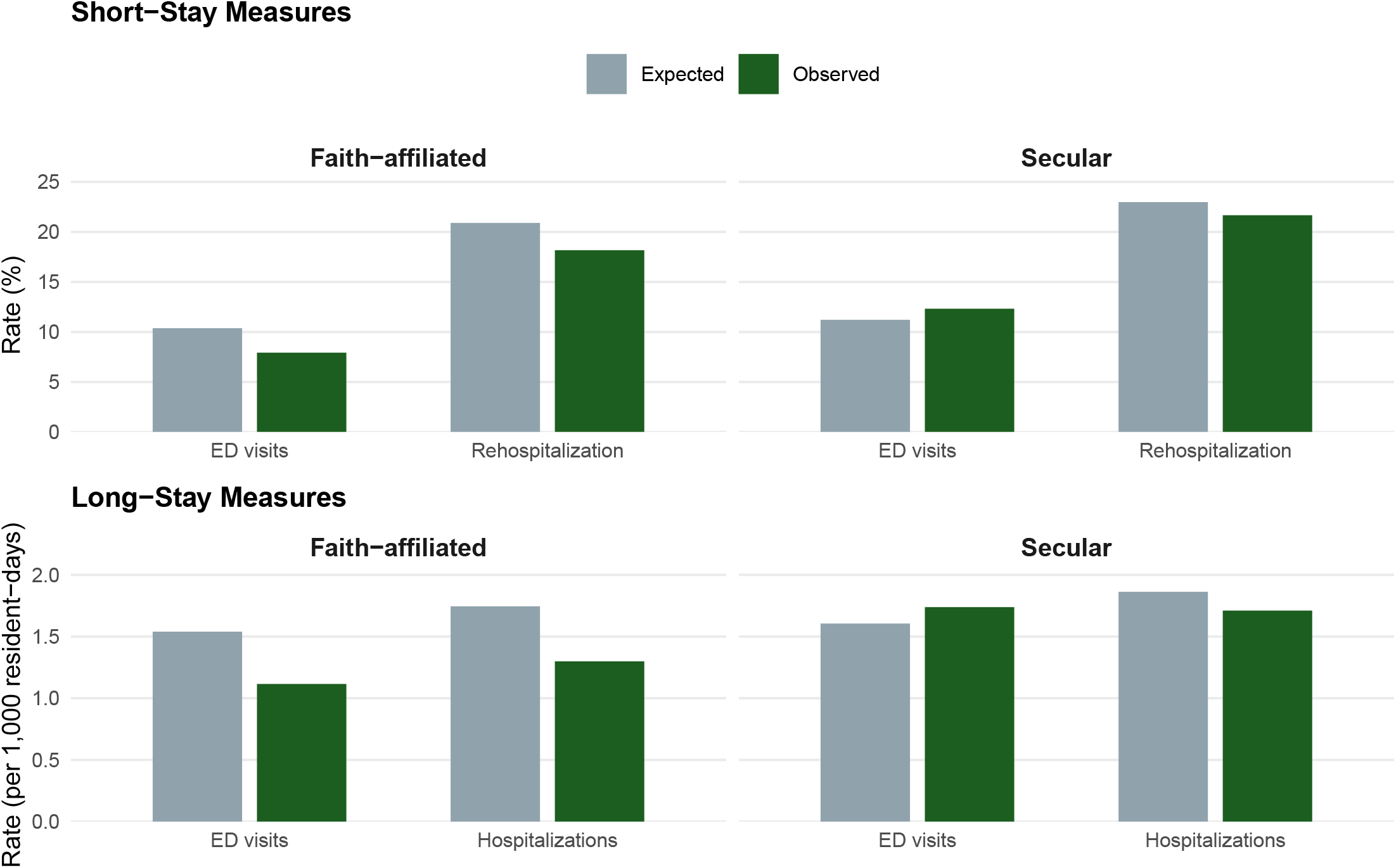
Observed vs. Expected Rates in Matched Sample. Observed vs. expected hospitalization and ED transfer rates in propensity-matched sample (n=49 matched pairs). Faith-affiliated facilities consistently performed below expected rates across all four measures, while matched secular facilities performed near expectations. Short-stay measures (521, 522) expressed as percentages; long-stay measures (551, 552) expressed as events per 1,000 resident-days.

### Mechanisms: From Organizational Identity to Staffing Intensity

The finding that faith affiliation predicts higher RN staffing is consistent with organizational theory literature on mission-driven institutions^10,23^. Faith-affiliated organizations may be more willing to sustain above-market RN staffing levels during periods of financial pressure, treat staffing as a mission-constrained rather than purely cost-minimizing decision, and attract RNs who self-select into environments aligned with their values^6^.

Drawing on Scott’s institutional pillars framework, faith identity creates normative pressure to staff at levels reflecting resident dignity and pastoral care commitments rather than margin optimization alone^10^. This normative pillar operates independently of regulatory compliance: faith facilities in our sample exceeded minimum staffing requirements not because regulations demanded it, but because organizational mission defined quality standards that surpassed regulatory floors. Additionally, Battilana and Dorado’s work on hybrid organizations suggests that faith facilities may use hiring-for-values strategies that attract and retain RNs who prioritize mission alignment over compensation, creating a self-reinforcing culture in which high-quality care is sustained through workforce selection as much as through wage premiums^23^. The observed 61% RN staffing advantage is too large to attribute to wage differences alone and more likely reflects both mission-driven investment decisions and values-based workforce attraction.

The substantial mediation by RN hours is particularly informative. It suggests that the mechanism linking organizational identity to hospitalization rates is not direct — faith affiliation does not appear to improve outcomes through, for example, family engagement or spiritual programming alone. Rather, the data are consistent with a model in which faith-driven values translate into staffing decisions that in turn improve clinical surveillance and early intervention. RNs, relative to LPNs and certified nursing assistants, are better positioned to identify early signs of clinical deterioration and initiate timely treatment, potentially preventing the cascade that leads to emergency transfer.

This interpretation is consistent with the broader literature on RN staffing and nursing home quality^24,25^. Our full-cohort analysis (n=9,752) confirms that the RN hours–composite gap relationship is strong and robust in the population: every additional RN hour per resident per day was associated with a 0.45 SD improvement in composite gap (p<10). The faith-affiliated staffing advantage (0.36 hours/resident/day) translates, through this coefficient, into a predicted gap improvement of 0.16 SD — closely matching the observed matched comparison of 0.38 SD, with the difference plausibly attributable to additional unmeasured cultural and operational factors.

### Consistency Across Faith Traditions

The finding that all 14 represented faith traditions showed negative mean composite gaps — and that none contributed to the significant underperformer category — strengthens the inference that the effect is associated with faith organizational identity broadly rather than with specific denominational practices or resources. This pattern argues against explanations based on, for example, access to denominational capital subsidies (which would predict Catholic or Lutheran advantages but not necessarily Baptist or Mennonite), and is more consistent with a cross-cutting organizational culture or mission-orientation mechanism.

Notably, the tradition with the strongest performance was Mennonite (mean gap = −1.39), followed by Presbyterian (−0.89) and Jewish (−0.85), with Catholic facilities (n=14, the largest faith subsample) showing a mean gap of −0.52. This within-tradition variation suggests that the faith advantage is not merely an artifact of any single well-resourced denomination but rather reflects shared commitments to mission-driven care that transcend specific theological traditions. The consistency across Protestant, Catholic, and Jewish facilities—traditions with distinct governance structures, funding models, and historical relationships to healthcare provision—points to organizational culture as the active ingredient rather than denomination-specific institutional supports.

### Policy Implications

These findings have several implications for nursing home quality policy and improvement.

First, they suggest that organizational culture is a meaningful upstream determinant of SNF quality, operating through its effects on staffing decisions. Current quality improvement interventions largely target facility-level operational practices — standardized protocols, QAPI processes, care transitions checklists. Our findings suggest that interventions targeting the organizational values and management philosophy that underlie staffing investment may be comparably important.

Second, the mediation by RN hours identifies a concrete, policy-actionable mechanism. Minimum nurse staffing standards were proposed and finalized in 2024 under the Biden administration, establishing the first federal minimum staffing requirements (0.55 RN hours per resident per day)^14^. This rule was subsequently repealed in December 2025 citing rural workforce challenges^15^. Our findings provide independent evidence that RN hours, specifically, are associated with reduced hospitalization rates at the facility level — complementing the individual resident-level evidence base used to justify minimum staffing policy. The faith-affiliated facilities in our sample averaged 0.96 RN hours per resident per day, 75% above the (now-repealed) federal minimum, suggesting that high-performing facilities already exceed regulatory floors by substantial margins.

Third, the cross-facility pattern documented here — that a subset of facilities consistently achieves lower-than-expected hospitalizations — is potentially actionable through payment design. Medicare Advantage plans and health systems operating under readmission penalty programs have direct financial incentives to identify and preferentially refer patients to high-performing facilities. Value-based payment models that make performance variation visible and reward sustained overperformance could accelerate diffusion of the organizational practices underlying it.

These facility-level findings create a foundation for resident-level risk prediction. Combining organizational culture indicators (faith affiliation, RN staffing intensity) with individual resident clinical trajectories could enable precision targeting of interventions to high-risk residents in low-staffing facilities—the segment most likely to benefit from increased clinical surveillance. Such predictive models represent the natural next step for translating these findings into actionable quality improvement tools. Integration of facility culture indicators with resident-level clinical trajectories, leveraging longitudinal EHR data and sequential decision frameworks developed for dynamic resource allocation, represents a promising direction for precision quality improvement in long-term care.

### Limitations

Several limitations warrant consideration. First, the cross-sectional design precludes causal inference about the organizational identity–performance relationship. While propensity score matching addresses observed confounding by facility size, ownership, and urbanicity, unmeasured confounders — including regional market concentration, hospital referral patterns, payer mix, and administrator tenure — may explain some of the observed performance differential.

Second, facility classification was performed by the research team using publicly available records, without access to resident-level or internal organizational data. While public records are generally reliable for identifying religious affiliation, misclassification is possible, particularly for facilities that have changed ownership, secularized over time, or have nominal rather than operational faith identities. We addressed this through a structured classification protocol, supplementary observations on ownership history and affiliation transitions, and formal inter-rater reliability assessment (kappa = 0.545, 94% initial agreement, 100% post-adjudication agreement across a 50-facility random sample^16^).

Third, the classified sample of 618 facilities, while substantially larger than prior studies of this question, represents 4.6% of facilities with computable performance data. The stratified sampling design prioritized tail facilities, which means the sample is not representative of the full distribution; gradient analyses should be interpreted with this sampling frame in mind.

Fourth, the composite gap measure, while a useful summary, aggregates across four measures with different denominators and clinical mechanisms. We addressed this limitation through robustness checks analyzing short-stay and long-stay composites separately, which confirmed consistent effects across care settings.

Finally, the mediation analysis follows the Baron-Kenny causal steps approach, which is subject to the standard limitations of causal mediation inference including potential mediator-outcome confounding and the assumption of no unmeasured confounding. Future work could employ more sophisticated mediation techniques including structural equation modeling or causal mediation frameworks that formally test mediation assumptions.

## Conclusions

Faith and mission-driven organizational identity is associated with substantially lower-than-expected hospitalization and emergency department transfer rates in a national stratified sample of skilled nursing facilities, an effect mediated by elevated RN staffing intensity. Specifically, faith-affiliated facilities achieved 3.5 percentage points fewer short-stay rehospitalizations and 0.41 fewer long-stay hospitalizations per 1,000 resident-days than matched secular facilities serving comparable patient populations. These findings are consistent across all 14 represented faith traditions, robust to propensity score matching on major structural confounders, and replicated across both short-stay rehabilitation and long-stay chronic care settings.

Organizational culture—operationalized here as faith-driven mission identity—predicts superior hospitalization performance through its effect on RN staffing intensity. This mechanism is actionable: facilities can invest in RN hours, and policymakers can incentivize such investment through minimum staffing standards and value-based payment design. Identifying high-performing facilities and understanding their organizational practices represents an immediately implementable strategy for improving nursing home quality at scale. The values that govern staffing investment decisions are modifiable upstream determinants of nursing home quality, and identifying the mechanisms linking mission identity to clinical outcomes represents a productive direction for quality improvement research and policy.

## Data Availability

All data are available online at: https://data.cms.gov/provider-data

## Data availability

All data used in this study are publicly available from the Centers for Medicare and Medicaid Services Provider Data Catalog (data.cms.gov/provider-data). Analysis code will be made available upon publication at the corresponding author’s GitHub repository.

## References

1. Mor V, Intrator O, Feng Z, Grabowski DC. The revolving door of rehospitalization from skilled nursing facilities. Health Affairs. 2010;29(1):57–64. doi:10.1377/hlthaff.2009.0629

2. Ouslander JG, Berenson RA. Reducing unnecessary hospitalizations of nursing home residents. New England Journal of Medicine. 2011;365(13):1165–1167. doi:10.1056/NEJMp1105449

3. Grabowski DC, O’Malley AJ, Barhydt NR. The costs and potential savings associated with nursing home hospitalizations. Health Affairs. 2007;26(6):1753–1761. doi:10.1377/hlthaff.26.6.1753

4. Intrator O, Grabowski DC, Zinn J, et al. Facility characteristics associated with hospitalization of nursing home residents: Results of a national study. Medical Care. 2007;45(10):921–930. doi:10.1097/MLR.0b013e3180616d79

5. Stevenson DG, Intrator O, Degenholtz HB, Zinn JS, Mor V. Predictors of nursing home and hospital use at the end of life among medicare beneficiaries. Journal of Palliative Medicine. 2013;16(4):360–367. doi:10.1089/jpm.2012.0330

6. Temkin-Greener H, Zheng NT, Katz P, Zhao H, Mukamel DB. Measuring work environment and performance in nursing homes. Medical Care. 2009;47(4):482–491. doi:10.1097/MLR.0b013e318194fbda

7. Konetzka RT, Yi D, Norton EC, Kilpatrick KE. Effects of medicare payment changes on nursing home staffing and deficiencies. Health Services Research. 2004;39(3):463–488. doi:10.1111/j.1475-6773.2004.00240.x

8. Harrington C, Woolhandler S, Mullan J, Carrillo H, Himmelstein DU. Does investor ownership of nursing homes compromise the quality of care? American Journal of Public Health. 2001;91(9):1452–1455. doi:10.2105/ajph.91.9.1452

9. Grabowski DC, Hirth RA. Competitive spillovers across non-profit and for-profit nursing homes. Journal of Health Economics. 2003;22(1):1–22. doi:10.1016/S0167-6296(02)00076-3

10. Scott WR. Institutions and Organizations: Ideas, Interests, and Identities. 4th ed. Sage Publications; 2014.

11. Comondore VR, Devereaux PJ, Zhou Q, et al. Quality of care in for-profit and not-for-profit nursing homes: Systematic review and meta-analysis. BMJ. 2009;339:b2732. doi:10.1136/bmj.b2732

12. Ryskina KL, Zhu JM, Unruh MA, Linn KA, Werner RM. Changes in nursing home quality following ownership conversions from nonprofit to for-profit. JAMA Health Forum. 2024;5(1):e234856. doi:10.1001/jamahealthforum.2023.4856

13. Li Y, Cai X, Mukamel DB, Glance LG. Nursing home quality after the COVID-19 pandemic: A systematic review. Journal of the American Medical Directors Association. 2023;24(8):1169–1178. doi:10.1016/j.jamda.2023.05.015

14. Centers for Medicare & Medicaid Services. Medicare and medicaid programs; minimum staffing standards for long-term care facilities and medicaid institutional payment transparency reporting. Published online 2024. https://www.federalregister.gov/documents/2024/05/10/2024-09766/medicare-and-medicaid-programs-minimum-staffing-standards-for-long-term-care-facilities-and

15. U.S. Department of Health and Human Services. HHS cleanup of federal nursing home minimum staffing standards rule expands access to rural, tribal health care. Published online 2025. https://www.hhs.gov/press-room/hhs-cleanup-federal-nursing-home-minimum-staffing-standards-rule-expands-access-rural-tribal-health-care.html

16. Landis JR, Koch GG. The measurement of observer agreement for categorical data. Biometrics. 1977;33(1):159–174. doi:10.2307/2529310

17. Baron RM, Kenny DA. The moderator-mediator variable distinction in social psychological research: Conceptual, strategic, and statistical considerations. Journal of Personality and Social Psychology. 1986;51(6):1173–1182. doi:10.1037/0022-3514.51.6.1173

18. R Core Team. R: A Language and Environment for Statistical Computing. R Foundation for Statistical Computing; 2025. https://www.R-project.org/

19. Wickham H, Averick M, Bryan J, et al. Welcome to the tidyverse. Journal of Open Source Software. 2019;4(43):1686. doi:10.21105/joss.01686

20. Ho DE, Imai K, King G, Stuart EA. MatchIt: Nonparametric preprocessing for parametric causal inference. Journal of Statistical Software. 2011;42(8):1–28. doi:10.18637/jss.v042.i08

21. Frick H, Chow F, Kuhn M, Mahoney M, Silge J, Wickham H. Rsample: General Resampling Infrastructure. 2023. https://CRAN.R-project.org/package=rsample

22. Robinson D, Hayes A, Couch S. Broom: Convert Statistical Objects into Tidy Tibbles. 2023. https://CRAN.R-project.org/package=broom

23. Battilana J, Dorado S. Building sustainable hybrid organizations: The case of commercial microfinance organizations. Academy of Management Journal. 2010;53(6):1419–1440. doi:10.5465/amj.2010.57318391

24. Harrington C, Ross L, Chapman S, Halifax E, Spurlock B, Bakerjian D. Nurse staffing and coronavirus infections in california nursing homes. Policy, Politics, & Nursing Practice. 2020;21(3):174–186. doi:10.1177/1527154420938707

25. Bostick JE, Rantz MJ, Flesner MK, Riggs CJ. Systematic review of studies of staffing and quality in nursing homes. Journal of the American Medical Directors Association. 2006;7(6):366–376. doi:10.1016/j.jamda.2006.01.024

